# Blood transfusion in care of patients with Visceral Leishmaniasis: a review of practices in therapeutic efficacy studies

**DOI:** 10.1101/2023.09.05.23295055

**Authors:** Prabin Dahal, Sauman Singh-Phulgenda, James Wilson, Glaucia Cota, Koert Ritmeijer, Ahmed Musa, Fabiana Alves, Kasia Stepniewska, Philippe J Guerin

## Abstract

Anaemia is a common presentation feature in patients with visceral leishmaniasis (VL). Blood transfusion remains an important aspect of patient management in VL. However, triggers considered for making decisions on transfusion are poorly understood. This review is based on the Infectious Diseases Data Observatory (IDDO) VL clinical trials library, a database of all published efficacy studies since 1980 and has indexed 160 published trials (1980–2021). Description of blood transfusion was reported in 16 (10.1%) trials (n=3,459 patients). Transfusion was initiated solely based on haemoglobin (Hb) measurement in 9 studies, using a combination of Hb and other health conditions (epistaxis, poor health, or clinical instability) in 3 studies, and the criteria was unclear in 4 studies; Hb threshold ranged from 3-8 g/dL. Overall, the number of patients receiving transfusion was explicitly reported in 10 trials (n=2,421 patients enrolled). Of these, 217 patients underwent transfusion; 58 before treatment initiation and 46 during treatment or the follow-up phase, and the time of transfusion was unclear in 113. The median proportion of patients who received a transfusion in a study was 8.0% [Interquartile range (IQR): 4.7% to 47.2%; range: 0-100%; n=10 studies]. This review describes the variation in current clinical practice and is an important initial step in policy/guideline development, where both the patient’s haemoglobin concentration and clinical status must be considered.

## Introduction

Visceral leishmaniasis (VL) is the most severe of the three forms of leishmaniasis and, is fatal without treatment. The disease typically presents insidiously and is characterised by protracted fever, hepatosplenomegaly and weight loss, often with evolving anaemia, leukopaenia and thrombocytopaenia leading to pancytopaenia.^1^ At presentation, moderate anaemia around 7–10 g/dL is common but can evolve into severe anaemia.^1^ While the underlying mechanism for the onset of anaemia in VL is likely multifactorial, the literature suggests the main cause is due to macrophage induced haemolysis in the spleen (“splenic sequestration, splenic haemophagocytosis”). ^2^

The WHO defines anaemia based on the haemoglobin levels below the reference values based on age, sex, and pregnancy status. ^3^ In patients presenting with severe anaemia, especially when accompanied by signs of anaemia (typically shortness of breath, fatigue, weakness, light-headedness, occasionally chest pain), transfusion of blood products may be therefore clinically indicated. Red blood cell (RBC) transfusion may also be indicated in cases of acute blood due to invasive procedures such as splenic aspiration or venous catheterisation or due to spontaneous haemorrhage from a VL-related bleeding diathesis. Bleeding risk is exacerbated in VL patients as the disease can lead to alterations of hepatic coagulation factors and thrombocytopaenia.^4^ Therefore, operational manuals from national control programmes suggest that haematological factors are taken into consideration prior to splenic aspirations (or completely contra-indicate splenic aspiration) and that blood transfusion services are in place for the management of acute blood loss. ^4–8^

Despite a clear recognition of the importance of transfusion in the management of VL patients, the actual transfusion practice in clinical trials or daily management of patients is poorly documented. In this review, we aim to collate information on blood transfusion among VL patients enrolled in therapeutic efficacy trials.

## Methodology

### Information sources and search strategy for clinical trials

This review synthesises data from studies indexed in the Infectious Diseases Data Observatory (IDDO) VL clinical trials library of prospective therapeutic studies from 1980 until Nov-2021 ^9^; details of the search strategy adopted for each of the databases have been described elsewhere.^10^ Data on the following aspects of design and conduct of studies captured by the IDDO VL library were extracted: the number of participants enrolled, details on the timing of transfusion (baseline, during treatment phase, or during the follow-up phase after discharge), and the details of the blood products transfused (whole blood, packed red blood cells (PRBC), plasma, or platelets), the haemoglobin threshold used for patient inclusion, and the transfusion triggers adopted.

### Data summary and analysis

A descriptive summary of the data extracted is presented. The median proportion of patients reported to have received blood products is presented along with the range and inter-quartile range. No formal meta-analysis was undertaken owing to the large heterogeneity in transfusion practices adopted. All statistical analyses were carried out using R software.^11^

### Risk of bias assessment

The risk of bias assessment in studies included in this review was carried out using the Cochrane Risk of Bias (ROB) tool for randomised controlled trials.^12^ Risk of bias in non- randomised studies will be carried out using ROBINS-I tool.^13^ Two reviewers (PD and SS) independently assessed the risk of bias.

## Results

The IDDO systematic review library has currently indexed 160 publications (35,763 patients; 1980–2021). There were 108 (67.5%) studies from the Indian subcontinent, 27 (16.9%) from East Africa, 9 (5.6%) from the Mediterranean region, 7 (4.4%) from South America, 5 (3.1%) from Central Asia (the Middle East), and 4 (2.5%) were multi-regional studies. The haematological measures adopted for patient inclusion are presented in Table 1. The minimum haemoglobin concentration required for inclusion was 3 g/dL in 8 (5.0%) studies, >3-5 g/dL in 38 (23.8%) studies, >5-7 g/dL in 22 (13.8%) studies, and unclear in the remaining 92 (57.5%) studies. Ranges of other heamatological parameters considered at inclusion are presented in Table 1.

**Table 1:** Haematological parameters considered for defining inclusion/exclusion criteria for patient enrolment.

| **Haematological parameters** | **Number of studies**<br>**(number of patients)** | **% (n=160)** |
| --- | --- | --- |
| Minimum haemoglobin concentration |  |  |
| >3 g/dL | 8 (n=1,804) | 5.0% |
| >3 to ≤5 g/dL | 38 (n=11,452) | 23.8% |
| >5 to ≤7 g/dL | 22 (n=2,644) | 13.8% |
| Unclear | 92 (n=19,863) | 57.5% |
| Prothrombin time (above control values) |  |  |
| >4 seconds | 9 (n=1,277) | 5.6% |
| >5 seconds | 14 (n=5,599) | 8.8% |
| >15 seconds | 2 (n=631) | 1.3% |
| >20 seconds | 1 (n=89) | 0.6% |
| Prothrombin activity <40% | 1 (n=57) | 0.6% |
| INR >2 | 1 (n=378) | 0.6% |
| Unclear | 132 (27,732) | 82.5% |
| Minimum platelets concentration |  |  |
| >4,000/μL | 1 (n=30) | 0.6% |
| >5,000/μL | 1 (n=60) | 0.6% |
| >20,000/μL | 1 (n=378) | 0.6% |
| >30,000/μL | 2 (n=38) | 1.3% |
| >40,000/μL | 22 (n=7,432) | 13.8% |
| >50,000/μL | 19 (2,681) | 11.9% |
| >60,000/μL | 1 (n=230) | 0.6% |
| >75,000/μL | 1 (n=120) | 0.6% |
| >80,000/μL | 4 (n=50) | 2.5% |
| Unclear | 108 (n=24,274) | 67.5% |
| Minimum WBC count |  |  |
| <750/μL | 1 (n=412) | 0.6% |
| <1,000/μL | 18 (n=7,086) | 11.3% |
| <2,000/μL | 8 (n=756) | 5.0% |
| <1,000/mL | 1 (n=30) | 0.6% |
| <2,000/nL | 1 (n=39) | 0.6% |
| granulocytes <1,000/μl | 18 (n=3,636) | 11.3% |
| granulocytes <2,000/μl | 1 (n=45) | 0.6% |
| Unclear | 112 (n=23,763) | 70.0% |

INR= International normalisation ratio

### Transfused products and transfusion triggers (n=16 studies)

Of the 160 studies in the IDDO VL systematic review library, the description of blood transfusion was explicitly reported in 16 (10.1%) studies (n=3,459 patients; 1984- 2018)(Table 2). 7/108 studies from the Indian sub-continent, 6/27 studies from East Africa, 2/7 studies from South America, 1/5 studies from Central Asia and none of the 9 studies from the Mediteeranean region or none of the 4 multi-regional studies reported data on blood transfusion. Patients living with VL-HIV co-infection were included in 2 studies (n=113 patients), excluded in 4 studies (n=384 patients), and unclear in the remaining 10 studies (n=2,962 patients). Pregnant women were included in 3 studies (n=996 patients), excluded in 4 studies (n=304 patients), and unclear in the remaining 9 studies (n=2,159 patients).

**Table 2:** Description of transfusion in clinical studies of VL indexed in the IDDO living systematic review library.

| Author-year | Country | Drug regimen | Total enrolled | Number of transfusions | Transfusion rules | Transfused product | Description | Number of Patients requiring transfusion at baseline | Transfusion during treatment /follow-up |
| --- | --- | --- | --- | --- | --- | --- | --- | --- | --- |
| Rees-1984<sup>21</sup> | Kenya | SSG | 16 | 1 | Unclear if there were any specific rules (the transfused patient had Hb measurement of 3.2 g/dL) | Blood | In one pregnant patient with a haemoglobin of 3.2 g/dL, 2 pints of blood were given concomitantly with the commencement of SSG | - | Transfused during treatment; unclear number of patients |
| Thakur-1984<sup>14</sup> | India | SSG | 750 | 4 | 4 g/dL +poor health OR haemorrhage | Blood | If Hb was below 4.0 g/dL and the general condition of the patient was poor, a blood transfusion was given. Blood transfusions were given to all cases with haemorrhage.<br>Four cases of cancrum oris were encountered in the early phase of the epidemic when drugs were scarce. With specific treatment, general management with blood transfusion and oral protein supplements and crystalline penicillin, cancrum oris improved. | - | - |
| Thakur-1988<sup>42</sup> | India | SSG | 371 | 0 | Unclear | No transfusion | As our patients were selected so that they did not have a haemoglobin concentration of less than 3 g/dL no blood transfusion was required for any patient | 0 | 0 |
| Thakur-1991<sup>43</sup> | India | Pentamidine; Pentamidine + SSG | 312 | Unclear | 3 g/dL | Blood | Patients whose haemoglobin level was less than 3 g/dL were given a blood transfusion, and when the haemoglobin level improved beyond 3 g/dL, they were included in the trial | Transfused at baseline; Unclear number of patients | - |
| Dietze-1993<sup>44</sup> | Brazil | ABCD | 20 | Unclear | Unclear | Red cells | Some patients were administered transfusions of red blood cells during therapy | - | Transfused during treatment; unclear number of patients |
| Thakur-1993<sup>23</sup> | India | Amphotericin B | 50 | 4 | 4 g/dL + pre-existing hemorrhagic problems | Blood | Patients were given blood transfusions if they had haemoglobin below 4 g/dL, or had any hemorrhagic problems like epistaxis. Epistaxis occurred in 4 patients and it was treated with blood transfusions (Table 1 of the publication indicates transfusions occurred before treatment as no epistaxis occurred) | 4 | 0 |
| Berhe-1999<sup>25</sup> | Ethiopia | PA | 23 | Unclear | severe anaemia | Blood | Blood transfusion was given to patients with severe anaemia and the post-transfusion haemoglobin value among the two groups was not different | - | - |
| Thakur-1999<sup>45</sup> | India | AMBd | 938 | 56 | 5 g/dL | Blood | The initial precaution taken was that patients whose haemoglobin level was below 5 g/dL were given blood transfusion first and only when the haemoglobin reached 5 g/dL was treatment for visceral leishmaniasis started. Blood transfusion was required in 54 (6%) patients before start of treatment. Anaemia improved with treatment but in 2 patients, haemoglobin dropped after treatment and they required blood transfusion | 54 | 2 |
| Moore-2001<sup>46</sup> | Kenya | SSG | 102 | 43 | anaemia | Blood | Blood transfusions were available for anaemic patients. Patients required transfusions during and after treatment; as described in Table 2 of the manuscript<sup>46</sup> | - | 43 |
| Haidar-2001<sup>24</sup> | Yemen | SSG | 32 | 24 | Unclear | Blood | Blood transfusions were required for 24 patients (73%) | - | - |
| Mueller-2008<sup>47</sup> | Uganda | AMBd; PA | 371 | Unclear | severe anaemia | Blood | Blood transfusions for severe anaemia were given, if necessary | - | - |
| Das-2009<sup>48</sup> | India | AMBd; Pentamidine | 82 | Unclear | 6 g/dL | Blood | In cases of VL, who had severe anaemia (Hb < 6 g/dl), before excluding such patients from the study, an effort was undertaken to increase their haemoglobin level by giving fresh blood transfusion as required | Transfused at baseline; Unclear number of patients | - |
| Adam-2009<sup>22</sup> | Sudan | SSG | 42 | 42 | severe anaemia | Blood | All 42 patients received blood transfusions for severe anaemia | - | - |
| Thakur-2010<sup>17</sup> | India | Amphotericin B | 230 | Unclear | 5 g/dL | Whole blood/platelets | If patients had haemoglobin levels < 5 g/dL or a thrombocyte count < 65,000 cells/μL, whole blood transfusions or platelet transfusions were given, respectively. If these two parameters reached acceptable levels, only then were the patients included in the trial | Transfused at baseline; Unclear number of patients | - |
| Cota-2014<sup>16</sup><br>(HIV negative) | Brazil | PA; AMBd; L-AmB | 46 | 19* | Measured<br>Hb/platelets/INR +<br>clinical<br>instability/bleeding | Red cells or<br>plasma or<br>platelets | In this study, all types of blood products were<br>accounted for between baseline and the end of<br>hospitalisation for VL treatment.<br>Specific transfusion rules:<br>platelets: <20,000 + active bleeding or below<br>50,000 before performing invasive procedures;<br>red cells: severe anaemia or Hb < 8 g/dL + clinical<br>instability;<br>plasma: INR > 1,5 + active bleeding | - | - |
| Cota-2014<sup>16</sup><br>(HIV positive) | Brazil | PA; AMBd; L-AmB | 44 | 23* | Measured<br>Hb/platelets/INR +<br>clinical<br>instability/bleeding | Red cells or<br>plasma or<br>platelets | In this study, all types of blood products were<br>accounted for between baseline and the end of<br>hospitalisation for VL treatment.<br>Specific transfusion rules:<br>platelets: <20,000 + active bleeding or below<br>50,000 before performing invasive procedures;<br>red cells: severe anaemia or Hb < 8 g/dL + clinical<br>instability;<br>plasma: INR > 1,5 + active bleeding | - | - |
| Mbui-2018<sup>20</sup> | Kenya,<br>Uganda | Miltefosine | 30 | 1 | Unclear | Unclear | The first had a case of “transfusion reaction” that<br>was considered an important medical event by<br>the investigator, occurring on day 203 after the<br>treatment start | - | 1 |

Three trials that reported provision for haematological profile correction prior to patients enrolment without describing if any transfusions were carried out; these studies are not included in this table<sup>15,40,41</sup>.
Hb= haemoglobin; SSG = Sodium stibogluconate; PA = Pentavalent antimony; AMB = Amphotericin B; AMBd = Amphotericin B deoxycholate; ABCD= Amphotericin B colloidal dispersion

Transfusion was not required for any patient in 1 study, and in the remaining 15 studies, transfusion of either whole blood or other blood products was reported (Table 2). Transfusion of blood (without specifying if whole blood was transfused or the blood components) was reported in 12 studies, 1 study transfused red blood cells, and 2 studies reported transfusing either blood (without making distinction if whole blood or packed RBC) or platelets.

Transfusion was initiated solely based on the measured concentration of haemoglobin (or anaemia status) or platelet concentrations in 9 studies (Table 2). In 2 studies from India, the transfusion trigger was a combination of measured haemoglobin concentration and existing of clinical conditions such as epistaxis or poor health ^14,15^. In a study from Brazil ^16^, the measured concentration of red cells or international normalisation ratio (INR) in addition to the clinical condition of the patient was adopted; transfusion of RBC was undertaken if a patient had severe anaemia (haemoglobin <8 g/dL) along with clinical instability and plasma transfusion was undertaken if the INR >1.5 and the patient had active bleeding. The criteria used for blood transfusion were not stated in the remaining 4 studies (Table 2). The haemoglobin threshold used as a transfusion trigger was 3 g/dL in 2 studies, 4 g/dL in 2 studies, 5 g/dL in 2 studies, 6 g/dL in 1 study, 8 g/dL in 1 study, anaemia or severe anaemia status (without reporting the haemoglobin threshold) in 3 studies, and the threshold was not clear in 1 study.

Of the 2 studies that reported on platelet transfusion, a threshold of <65,000 cells/μL was used in a study from India ^17^, and in another study from Brazil [14], a threshold of <50,000 cells/μL was adopted among those who were given prophylactic transfusion before undertaking invasive procedures and a threshold of <20,000 cells/μL for patients with active bleeding (Table 2).

### The number of blood transfusions reported (n=10 studies)

The number of patients who received transfusions were clearly reported in only 10 trials (2,421 patients enrolled); a total of 217 patients received blood transfusions (total number of transfusion episodes were not reported). Of these 217 patients, 58 transfusions occurred before initiation of antileishmanial therapies ^15,18^, 46 patients underwent transfusion during treatment or follow-up phase ^18–20^, and the time when transfusion was carried out was not reported for the remaining 113 patients (Table 2). Overall, from the 10 studies that clearly reported transfusion data, the median proportion of patients who received a transfusion at any time-point in the study was 8.0% [Interquartile range (IQR): 4.7% to 47.2%; range: 0-100%] (Figure 1).

**Figure 1:**
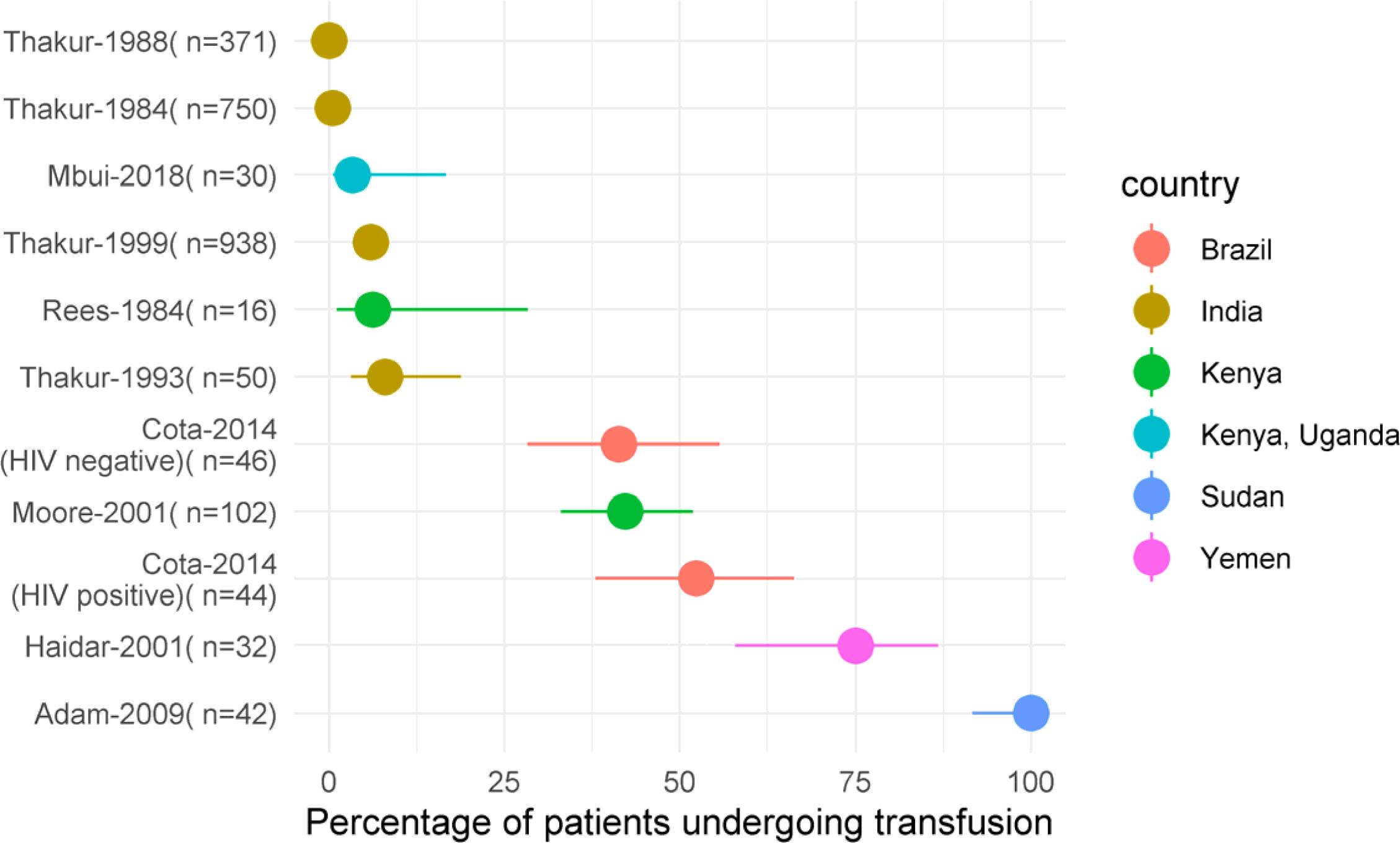
Proportion of patients undergoing transfusion in 10 studies that clearly reported the transfusion status

Two studies described transfusion among pregnant women: in a patient (1/16) in a study from Kenya (transfused at 3.2 g/dL) ^21^ and all patients (100%, 42/42) in another study on VL in pregnant women from Sudan (transfusion based on severe anaemia) ^22^. In 3 studies that explicitly enrolled children <15 years, of the 112 enrolled, 29 required transfusions (Table 2 and supplemental table 1); the criteria for transfusion was Hb concentration <4 g/dL along pre-existing hemorrhagic problems in a study from India ^23^ and this unclear in studies from Yemen ^24^ and East Africa ^20^.

Two studies enrolled patients with HIV co-infections; 23/44 (52.2%) patients from a study in Brazil ^16^ required transfusion; and the actual number of patients requiring transfusion was not reported in a study from Ethiopia (n=23 enrolled; number requiring transfusions was not reported) ^25^.

### Risk of bias assessment in studies included

Of the 16 studies included, 4 were randomised, and 12 were non-randomised studies. The four randomised studies were judged to be at high/unclear risk of bias on blinding domain, low or unclear risk of bias on sequence generation and allocation conceal domain. Of the 12 non-randomised studies, all of them were either open label or this description was unclear, bias in participants selection was considered low/moderate in 10 studies, high in 1 study, and unclear in 1 study. Risk of bias assessment is presented in supplemental file 1.

## Discussion

From the studies indexed in the IDDO VL clinical trials library, reporting of information on blood transfusion was not explicit in the majority of clinical trials. These could be partly due to the exclusion of patients with severe anaemia or severe disease in standard efficacy studies in VL. For example, approximately 40% of the 160 therapeutic efficacy studies excluded patients with haemoglobin concentrations less than 5 g/dL. The combination of inclusion/exclusion criteria adopted will likely lead to the exclusion of severe patients who may be less likely to require transfusion. From the 16 studies that clearly reported occurrences (or abscences) of blood transfusion, the criteria adopted for transfusion varied between the studies. Most of the studies reported initiating blood transfusion based on thresholds of haemoglobin concentration or anaemia status, without taking other factors (for example, clinical stability/bleeding conditions/poor health) into consideration; the haemoglobin concentration used as a transfusion trigger ranged from 3-8 g/dL. This wide variation is particularly relevant as recent studies (non-VL context) have pointed towards the lack of benefit of blood transfusion in preventing mortality when transfusion is initiated at haemoglobin concentration greater than 4 g/dL.^26–29^ Such assessment of the risk-benefit ratio associated with transfusion was not possible in this review as clinical outcomes were not disaggregated by transfusion status in the included studies. In addition to these criteria, it is also important to consider further haemodynamic stability and clinical history of patients, such as heart conditions, when considering the adoption of a transfusion threshold.^30^

The current WHO guidelines on the management of paediatric anaemia recommend carrying out transfusion when haemoglobin concentration is below 4 g/dL, and among those with non-severe anaemia (haemoglobin 4-6 g/dL), transfusion is only indicated if the child presents with other clinical features including dehydration and heart failure.^31^ For adults, specific guidelines among critically ill patients in the ICU have advocated a threshold of 7 g/dL and have recommended further personalising the transfusion decision based on the clinical condition of the patient.^32^ However, only three studies included in this review reported carrying out transfusion using a combination of haemoglobin concentration and further clinical criteria such as clinical stability, poor health or active bleeding, such as epistaxis (Table 1). This is particularly relevant as the ability to tolerate anaemia can partly depend on the speed of its evolution, as compensatory mechanisms can enable relatively severe degrees of anaemia to be tolerated if it develops over a prolonged duration.^33^ VL primarily affects the poor and marginalised populations with limited access to healthcare, leading to a prolonged duration of illness prior to presentation. As the disease itself often evolves insidiously over weeks or months, and patients often receive care late in the disease course, anaemia evolves over a prolonged.^34^ Therefore, in VL patients, it can be anticipated that compensatory mechanisms will have led to a physiological adaptation to anaemia. However, acute anaemia arising as a result of acute bleeding occurring due to complications during splenic puncture or post-partum haemarroage among pregnant VL patients ^35^ can overwhelm the compensatory mechanisms of the body and can be fatal, thus requiring immediate transfusion.

Further caution is urged when transfusing patients who present with severe acute malnutrition (a common feature of VL patients), as fluid overload and respiratory impairment are a recognised and feared complication in patients with hypoalbuminemia.^31^ In the studies included in this review, one case of transfusion reaction was reported ^20^; but specific details on the nature of this adverse event were not presented. In other studies included in this review, no reports on the occurrence of transfusion reactions or transfusion-associated risks were reported. However, the absence of reporting of such occurrences cannot be taken as evidence of absence of adverse events. In general, transfusion reactions are estimated to occur up to 1 per 100 transfusions.^36^ There are no such estimates specific to the VL context, and the risks associated with transfusion in the context of VL are not well understood.

From the relatively limited set of studies that reported the occurrences of transfusions, the median proportion of patients who received blood transfusions was 8% (n=10 studies). As mentioned earlier, patients with severe anaemia or those with severe VL and pre-existing co-morbidities are excluded in standard therapeutic efficacy studies leading to the inclusion of mostly uncomplicated VL cases. Therefore, the incidence of transfusion in routine clinical practice is likely to be much higher. This suggests that there might be a substantial economic/logistic cost to the healthcare facilities arising from the requirement of transfusion in the management of VL patients. The cost could also be further increased as a central cause of anaemia may require multiple transfusions leading to increased expenditure and the management of potential safety risks associated with transfusion alone ^37–39^, while the benefit for the patients may not be warranted.

There are several limitations with this review. Of the 160 clinical studies in the VL IDDO systematic review library, the majority of the studies didn’t report any transfusion related data. Data on the number of transfusions carried out were also not reported; therefore, the actual number of transfusion episodes remains unclear. Overall, these indicate generally high or unclear risk of bias in the studies included with respect to assessing blood transfusion and their effect on treatment. However, since blood transfusion isn’t the main focus of the efficacy studies, lack of details regarding transfusion and outcomes preveted a thorough assessment of risk of bias. Instead, the overall quality of studies were assessed using standard risk of bias tools. Another aspect to be considered is the influence of the conditions of the health services where the patients gathered here were originally treated, including the possibility of different local guidelines for transfusions. For example, three studies reported undertaking haematological profile correction prior to enrolment of patients without describing if any transfusions were carried out.^15,40,41^ Such practice can affect the requirement for transfusion during the study. Similarly, access to blood products is relatively difficult in some countries, which may have contributed to the observed differences in the rate of transfusions between studies. Finally, from the reports included in this review, it was not possible to reliably assess if some patient groups were more or less likely to require transfusion than others; this would require an individual participant data meta-analysis.

A checklist of items for reporting data related to transfusion is proposed in Box 1. Adoption of such a checklist can facilitate better reporting of the transfusion-related parameters and can enable a thorough assessment of the risks and benefits of transfusion strategies adopted among VL patients in the future. Additionally, the standardisation and completeness of haematological data in VL studies before, during and after treatment, including information concerning blood transfusions, therapy and clinical outcome stratification, may also help in recognising the dynamics of VL clinical improvement after treatment, over time, contributing to the establishment of operational definitions to support cure assessment.

## Conclusions

Data regarding blood transfusions remain largely unreported in therapeutic efficacy studies on VL, with information available only on 16 therapeutic efficacy studies published since 1980. When reported, the decision to undertake transfusion was often found to be solely based on the haemoglobin concentration of the patients, with only three studies incorporating additional clinical criteria. Overall, this review represents the initial step in acknowledging the magnitude of the gap related to blood derivatives use and points to the need for harmonisation of the clinical data presentation in VL prospective studies. The research community should adopt a standardised method for the reporting oftransfusion episodes so that the true benefit of transfusion in VL case management can be reliably assessed.

## Authors’ contributions

Study Conception: PD, SSP, KS, PJG Methodology: PD, SSP

Data curation: PD, SSP

Project supervision: SSP, KS, PJG Project administration: PD, SSP Funding acquisition: PJG

Writing-original draft: PD, SSP, JW, GC, KR, AM, FA, KS, PJG

Writing- review and editing: All authors were involved in reading and critical revision of the initial draft and approved the final manuscript.

## Funding

This work is funded by a Bill & Melinda Gates Foundation grant to the Infectious Diseases Data Observatory, Oxford University, UK (Recipient: Prof. Philippe Guerin; ref: INV-004713). Funding agency had no role in developing the manuscript or its publication.

## Conflict of Interest

None

## Data Availability

All the data used in this review are available within supplemental file 1.

---

**Box 1:**
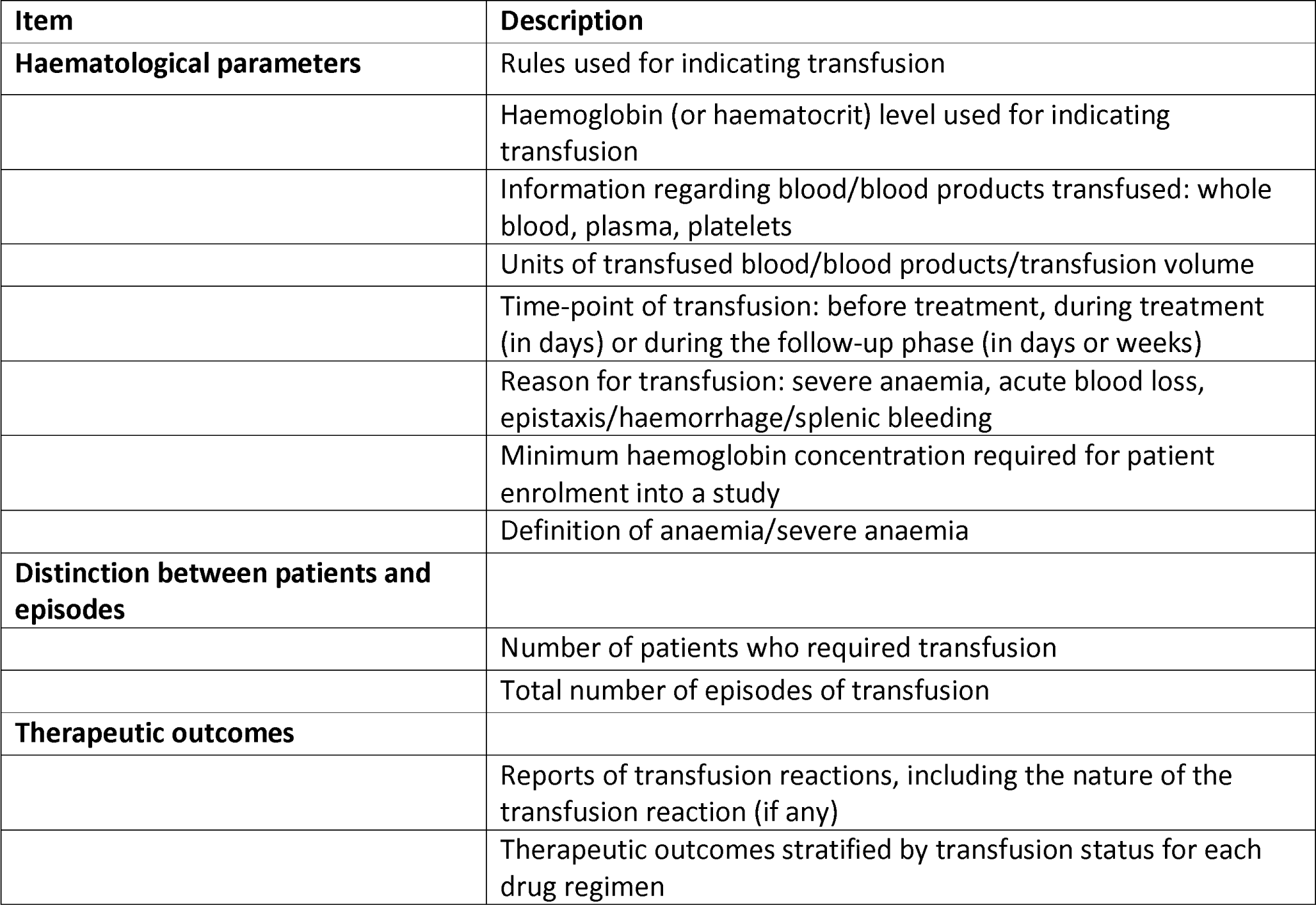
Transfusion parameters

## Notes

### Competing Interest Statement

The authors have declared no competing interest.

