## Supplemental File 1 for "Blood transfusion in care of patients with Visceral Leishmaniasis: a review of practices in therapeutic efficacy studies"

Blood transfusion in care of patients with Visceral Leishmaniasis: a review of practices in therapeutic efficacy studies

Prabin Dahal*^1,2^, Sauman Singh-Phulgenda^1,2^, James Wilson^1,2^, Glaucia Cota^3^, Koert Ritmeijer^4^, Ahmed Musa^5^, Fabiana Alves^6^, Kasia Stepniewska^1,2^, Philippe J Guerin*^1,2^

^1^Infectious Diseases Data Observatory (IDDO), Oxford, UK

^2^Centre for Tropical Medicine and Global Health, Nuffield Department of Medicine,

University of Oxford, Oxford, UK

^3^Instituto René Rachou (IRR), Fiocruz, Minas Gerais, Brazil

^4^Médecins Sans Frontières, Amsterdam, Netherlands

^5^Institute of Endemic Diseases, University of Khartoum, Khartoum, Sudan

^6^Drugs for Neglected Diseases initiative, Geneva, Switzerland

### Supplemental Table 1: Description of studies included in the review

| **Author-year** | **Country** | **Total enrolled** | **Duration of illness (days)** | **Age-range of included participants** | **Inclusion of HIV co-infected patients** | **Inclusion of pregnant women** | **Drug regimens**  **used for treatment** |
| --- | --- | --- | --- | --- | --- | --- | --- |
| Rees-1984 ^1^ | Kenya | 16 | 53.2 | All ages | Unclear | Included | SSG |
| Thakur-1984 ^2^ | India | 750 | - | All ages | Unclear | Unclear | SSG |
| Thakur-1988 ^3^ | India | 371 | 166.5 | All ages  (9-45y) | Unclear | Unclear | SSG |
| Thakur-1991 ^4^ | India | 312 | - | All ages  (2-60y) | Unclear | Unclear | Pentamidine; Pentamidine + SSG |
| Dietze-1993 ^5^ | Brazil | 20 | 90 | All ages  (1-57y) | Unclear | Unclear | ABCD |
| Thakur-1993 ^6^ | India | 50 | - | Children  (Less than 15y) | Unclear | Unclear | Amphotericin B |
| Berhe-1999 ^7^ | Ethiopia | 23 | - | Adults  (20-47y) | Included | Unclear | PA |
| Thakur-1999^8^ | India | 938 | - | All ages  (0.8-80y) | Unclear | Included | AMBd |
| Moore-2001^9^ | Kenya | 102 | - | All ages  (2-40y) | Unclear | Excluded | SSG |
| Haidar-2001 ^10^ | Yemen | 32 | 168.7 | Children  (<12y) | Unclear | Unclear | SSG |
| Mueller-2008 ^11^ | Uganda | 371 | 28-53 | All ages | Unclear | Unclear | AMBd; PA |
| Das-2009 ^12^ | India | 82 | - | All ages  (6-60y) | Excluded | Excluded | AMBd; Pentamidine |
| Adam-2009^13^ | Sudan | 42 | - | All ages  (29.7±14.5 y) | Excluded | Included | SSG |
| Thakur-2010 ^14^ | India | 230 | - | All ages  (5-55y) | Excluded | Unclear | Amphotericin B |
| Cota-2014 ^15^ (HIV negative) | Brazil | 46 | 60 | All ages  (37.1 ± SD: 14.0y) | Excluded | Excluded | PA; AMBd; L-AmB |
| Cota-2014 ^15^ (HIV positive) | Brazil | 44 | 60 | All ages  (41.0 ± SD: 10.9) | Included | Excluded | PA; AMBd; L-AmB |
| Mbui-2018 ^16^ | Kenya, Uganda | 30 | - | Children  (4-12y) | Excluded | Excluded | Miltefosine |

SSG = Sodium stibogluconate; PA = Pentavalent antimony; AMB = Amphotericin B; AMBd = Amphotericin B deoxycholate; ABCD= AMB = Amphotericin B colloidal dispersion; SD= standard deviation

### Supplemental Table 2: Risk of bias in randomised studies

| IDDO ID | Author-year | Blinding details | Study conduct details | Domain | | | | | |
| --- | --- | --- | --- | --- | --- | --- | --- | --- | --- |
|  |  |  |  | Random sequence  generation | Allocation  concealment | Blinding of  participants  and personnel | Blinding of  outcome assessment | Incomplete  outcome data addressed | Selective  reporting |
| 23 | Das-2009 | Open | Treatment allocation was done by the biostatistician of the  institute, who performed the allocation sequence using random number tables and accordingly assigned the test and control group. The patients were sent to the indoor ward for further treatment with their allotted drug. Both groups of patients were treated after the hospitalization in RMRIMS indoor ward. Every patient completed the full course of assigned treatment. | Low | Unclear | High | High | Low | Low |
| 39 | Thakur-1991b | Unclear | Patients were randomly allocated to three treatment groups. | Unclear | Unclear | Unclear | Unclear | Low | Low |
| 55 | Thakur-2010 | Open | This study was conducted as an open-label, randomized trial of 230 patients at Balaji Utthan Sansthan, Patna. The study staff who treated the patients opened consecutively numbered envelopes containing the treatment assignment after eligible patients fulfilled the entry criteria. Clinicians who provided treatment were not blinded to the treatment given. | Unclear | Low | High | Unclear | Low | Low |
| 70 | Thakur-1988 | Unclear | The patients were randomly allocated to six treatment groups. | Unclear | Unclear | Unclear | Low | Low | Low |

### Supplemental Table 3: Risk of bias in non-randomised studies

| **IDDO ID** | **Author-year** | **Description of study design and conduct** | **Bias due to confounding (Imbalances in baseline distribution)** | **Bias in selection  of participants** | **Bias in intervention  classification** | **Missing  outcome data** | **Bias in  outcome assessment** | **Selective  outcome reporting** | **Treatment**  **Blinding** |
| --- | --- | --- | --- | --- | --- | --- | --- | --- | --- |
| 9 | Berhe-1999 | Twenty-three consecutive HIV-VL patients with no other obvious concurrent infectious diseases were recruited from an ongoing VL-HIV co-infection study. | Not applicable;  single-armed trial | Low/moderate  (consecutive patient case series) | - | Unclear | Low | Unclear | Unclear |
| 31 | Thakur-1999 | Confirmed cases of visceral leishmaniasis were included in this study of all consecutive cases coming for treatment between 1 January and 31 December 1997 at the Kala-azar Research Centre of Balaji Utthan Sansthan, Patna. | Not applicable;  single-armed trial | Low/moderate  (consecutive patient case series) | - | Low | Low | Low | Unclear |
| 36 | Moore-2001 | The allocation of patients to treatment groups was alternate and not random, and the hospital staff were not blinded to the treatment given. However, the slides of splenic aspirates were read ‘‘blind’’, and the main outcome measures (death, initial cure, or definitive cure) are unlikely to have been affected by a knowledge of the treatment received. | Moderate/High (Table 1 of the manuscript) | Low/moderate (Table 1 of the manuscript) | Low | High | Low | Low | Open |
| 71 | Mueller-2008 | Between September 2003 and April 2004, the supply of antimonial drugs to Amudat Hospital, in north–eastern Uganda, was interrupted and all cases of visceral leishmaniasis presenting at the hospital could only be treated with amphotericin B deoxycholate (AmB). For comparison with the results of the AmB treatment, an historical cohort, of all the patients diagnosed with first-time VL when they presented at the Amudat Hospital between September 2002 and April 2003, was selected. The patients in this cohort had all been treated with intramuscular injections of SbV, given at 20 mg/kg.day (without an upper limit) for 30 days | Moderate/High (Table 1 of the manuscript gives reasonably similar distribution of the baseline covariates) | Low/moderate | Low | Low | Low | Low | Unclear |
| 82 | Dietze-1993 | This study was an open-label, phase 1/2 clinical trial of the efficacy and toxicity of Amphocil. Two cohorts, each consisting of 10 consecutive patients with kala-azar, were treated with Amphocil at a dosage of 2 mg kg/day. | Moderate/High (Table 1 of the manuscript gives reasonably similar distribution of the baseline covariates) | Low/moderate | Low | Low | Low | Low | Open |
| 103 | Thakur-1993a | Study in fifty children suffering from multiple drug resistant kala-azar, with classical features of severe kala-azar | Not applicable;  single-armed trial | High  (drug resistant VL cases enrolled in the study) | - | Low | Low | Low | Unclear |
| 111 | Haidar-2001 | A prospective hospital-based study in children of 12 years of age or less | Not applicable;  single-armed trial | Low/moderate | - | Unclear | Low | Unclear | Unclear |
| 117 | Rees-1984 | Sixteen consecutive new patients with kala-azar seen by us between September 1978 and February 1979 at the Kenyatta National Hospital, Nairobi, were studied | Not applicable;  single-armed trial | Low/moderate  (consecutive patient case series) | - | Unclear | Low | Low | Unclear |
| 127 | Thakur-1984 | Data from outbreak | Not applicable;  single-armed trial | Low/moderate as all data from the outbreak is reported | Low | Low | Low | Low | Unclear |
| 132 | Cota-2014 | The study was conducted at a reference centre for infectious diseases in Brazil. All patients with suspected VL were evaluated in an ongoing cohort study | Unclear  (table 1 stratified by HIV status) | Unclear | Low | Low | Low | Low | Unclear |
| 135 | Adam-2009 | Prospective cohort study | Not applicable;  single-armed trial | Low/moderate | - | Unclear | Low | Moderate | Unclear |
| 149 | Mbui-2018 | Open-label clinical trial | Not applicable;  single-armed trial | Low/moderate | - | Low | Low | Low | Open |
